# Cost-effectiveness of Evolocumab in Patients at High Cardiovascular Risk Without a Previous Myocardial Infarction or Stroke

**DOI:** 10.64898/2026.07.23.26358791

**Authors:** Gregg C. Fonarow, Cameron Cook, Eduard Sidelnikov, Sushmitha Inguva, Ajay Bhatia, Guillermo Villa

**Affiliations:** Ahmanson-UCLA Cardiomyopathy Center, Division of Cardiology, University of California Los Angeles, Los Angeles, CA, USA; Amgen Inc., Thousand Oaks, CA, USA; Amgen (Europe) GmbH, Rotkreuz, Switzerland

**Keywords:** evolocumab, cost-effectiveness, VESALIUS-CV, QALY, cardiovascular

## Abstract

**Importance:** Evolocumab reduces major adverse cardiovascular events (MACE) in patients with clinically evident atherosclerotic cardiovascular disease (ASCVD) and in patients at high cardiovascular (CV) risk but without a prior myocardial infarction (MI) or stroke. While evolocumab is shown to be cost-effective in clinically evident ASCVD, emerging CV outcomes evidence and newer guideline recommendations warrant assessment in patients at high CV risk without a prior MI or stroke.

**Objective:** To evaluate the cost-effectiveness of evolocumab added to standard therapy compared with standard therapy alone in patients at high CV risk without a prior MI or stroke, as represented by the VESALIUS-CV trial.

**Design, Setting, and Participants:** A previously published Markov cohort state-transition model was adapted to simulate VESALIUS-CV patients over a lifetime horizon. Health states included high-risk without a prior MI or ischemic stroke (IS), non-fatal MI, non-fatal IS, post-MI, post-IS, CV death, and non-CV death. Revascularization (RV) was modeled as a procedure with associated costs. The base case considered a US payer perspective and CV risk reduction inputs from evolocumab CV outcomes trials, including VESALIUS-CV, FOURIER, and FOURIER-OLE. Three scenario analyses were evaluated. Scenario 1 retained the US payer perspective and applied CV risk reduction estimates based on the Cholesterol Treatment Trialists’ (CTT) Collaboration 2010 meta-analysis. Scenarios 2 and 3 adopted a US societal perspective, using evolocumab CV outcomes trial–based and 2010 CTT Collaboration-based risk reduction estimates, respectively.

**Main Outcomes and Measures:** The model outcomes included MACE (defined as MI, IS, or CV death), RV procedures, total costs, life-years (LYs), quality-adjusted life-years (QALYs), and incremental cost-effectiveness ratio (ICER).

**Results:** In the base case, at the current direct-to-patient price of $3,107 per year, evolocumab added to standard therapy was associated with lifetime reductions of 0.24 MACE and 0.17 RV procedures per person, incremental costs of $24,430, incremental QALYs of 0.34, and an ICER of $71,162 per QALY gained. Evolocumab remained cost-effective across all evaluated scenarios, with ICERs of $42,094, $51,160, and $20,152 per QALY in Scenarios 1, 2, and 3, respectively.

**Conclusions and Relevance:** In patients at high CV risk without a prior MI or stroke, evolocumab added to standard therapy was projected to improve CV outcomes and quality-adjusted survival. At the current direct-to-patient price of $3,107 per year, evolocumab was cost-effective in the base-case analysis, with an ICER of $71,162 per QALY gained, and remained cost-effective across scenario analyses, with ICERs ranging from $20,152 to $51,160 per QALY. These estimates were substantially below the $120,000 per QALY threshold defined in the 2025 American Heart Association/American College of Cardiology cost/value methodology statement.

**Key Points:** *Question:* What is the cost-effectiveness of evolocumab in patients at high cardiovascular (CV) risk without a previous myocardial infarction (MI) or stroke, as represented in the VESALIUS-CV trial?

*Findings:* In this study using a Markov cohort state-transition model, evolocumab added to standard therapy compared with standard therapy alone was associated with an incremental cost-effectiveness ratio (ICER) of $71,162 per quality-adjusted life-year (QALY) gained, at the direct-to-patient price of $3,107 per year. In scenario analyses evaluating alternative CV risk reduction estimates and/or societal perspective, ICERs ranged from $20,152 to $51,160 per QALY at this price. Across all analyses, ICERs were substantially below the American Heart Association/American College of Cardiology cost-effectiveness threshold of $120,000 per QALY gained.

*Meaning:* For a high CV risk population without a previous MI or stroke, evolocumab added to standard therapy was projected to improve outcomes and be cost-effective across multiple modeled scenarios.

## Introduction

Elevated low-density lipoprotein cholesterol (LDL-C) is a key modifiable risk factor for atherosclerotic cardiovascular disease (ASCVD). In the US, ASCVD impacts about 26 million individuals, and many others have risk factors, including elevated LDL-C, that are associated with increased risk for developing incident ASCVD events.^1–3^ These populations contribute to substantial clinical and economic burden through high rates of hospitalization and deaths annually.^1–3^ In 2021 to 2022, the annual direct and indirect costs of cardiovascular (CV) disease were estimated at $414.7 billion in the US, with direct costs accounting for nearly 10% of the total US healthcare expenditures.^2^ Despite this burden, many patients remain undertreated, with real-world evidence showing persistent gaps in lipid management that leave them vulnerable to experiencing major CV events.^4–7^

Evolocumab is a fully human proprotein convertase subtilisin/kexin type 9 (PCSK9) monoclonal antibody that produces substantial reductions in LDL-C and has demonstrated CV outcome benefits in a broad spectrum of patients with elevated CV risk.^8^ In the FOURIER trial (NCT01764633), evolocumab reduced the risk of major CV events compared to placebo in patients with clinically evident ASCVD, defined as a history of myocardial infarction (MI), nonhemorrhagic stroke, or symptomatic peripheral artery disease.^9^ FOURIER-OLE (NCT03080935), the open-label extension of FOURIER, provided long-term data on safety and CV outcomes with evolocumab over a median follow-up of 5 years.^10^ More recently, the VESALIUS-CV trial (NCT03872401) extended evidence of CV benefit to patients with atherosclerosis or high-risk diabetes who had no prior MI or stroke.^11^ Over a median follow-up of 4.6 years, evolocumab demonstrated a 25% lower risk of first major CV events compared with placebo.^11^

Current American College of Cardiology (ACC)/American Heart Association (AHA) Multisociety Dyslipidemia guidelines recommend PCSK9 monoclonal antibodies for secondary prevention in adults with clinical ASCVD and for primary prevention in select high-risk groups, when LDL-C and/or non–high-density lipoprotein cholesterol remain above recommended goals despite statin-based lipid-lowering therapy.^12^ Given the evolving clinical evidence base and the recent guideline recommendations related to primary prevention, understanding the economic value of evolocumab in the broader patient population is needed.

The cost-effectiveness of evolocumab has been previously evaluated using data from the FOURIER trial.^13^ Following a list price reduction to $5,850 per year in 2018, a cost-effectiveness analysis of patients with very–high-risk ASCVD estimated incremental cost-effectiveness ratios (ICERs) of $7,667 to $56,655 per quality-adjusted life-year (QALY) gained from a US societal perspective, depending on the modeled baseline CV event rate.^14^ Referencing this cost-effectiveness analysis, the current ACC/AHA Multisociety Dyslipidemia guidelines subsequently characterized evolocumab as cost-effective for secondary ASCVD prevention.^12^ The current study adapted this same model to evaluate the cost-effectiveness of evolocumab added to standard therapy (statins with or without ezetimibe) compared with standard therapy alone based on the VESALIUS-CV population of patients at high CV risk without prior MI or stroke, using contemporary US model inputs.

## Methods

An adapted Markov cohort state-transition model was used with a lifetime horizon, and model health states aligned with the VESALIUS-CV trial.^11,14^ Health states included high-risk without a prior MI or ischemic stroke (IS), non-fatal MI, non-fatal IS, post-MI, post-IS, CV death, and non-CV death (Figure 1). Patients entered the model in the high-risk without a prior MI or IS health state, reflecting the VESALIUS-CV population of patients with atherosclerosis or high-risk diabetes and no prior MI or stroke, and transitioned to other states with specific probabilities. The model was used to predict first and subsequent CV events and mortality (either CV-related or non–CV-related). Revascularization (RV) was modeled as a procedure with associated costs. Given that age, recent event history, and history of multiple events influence the risk of experiencing an event, transition probabilities in the model were adjusted to reflect these factors.^15^

**Figure 1.**
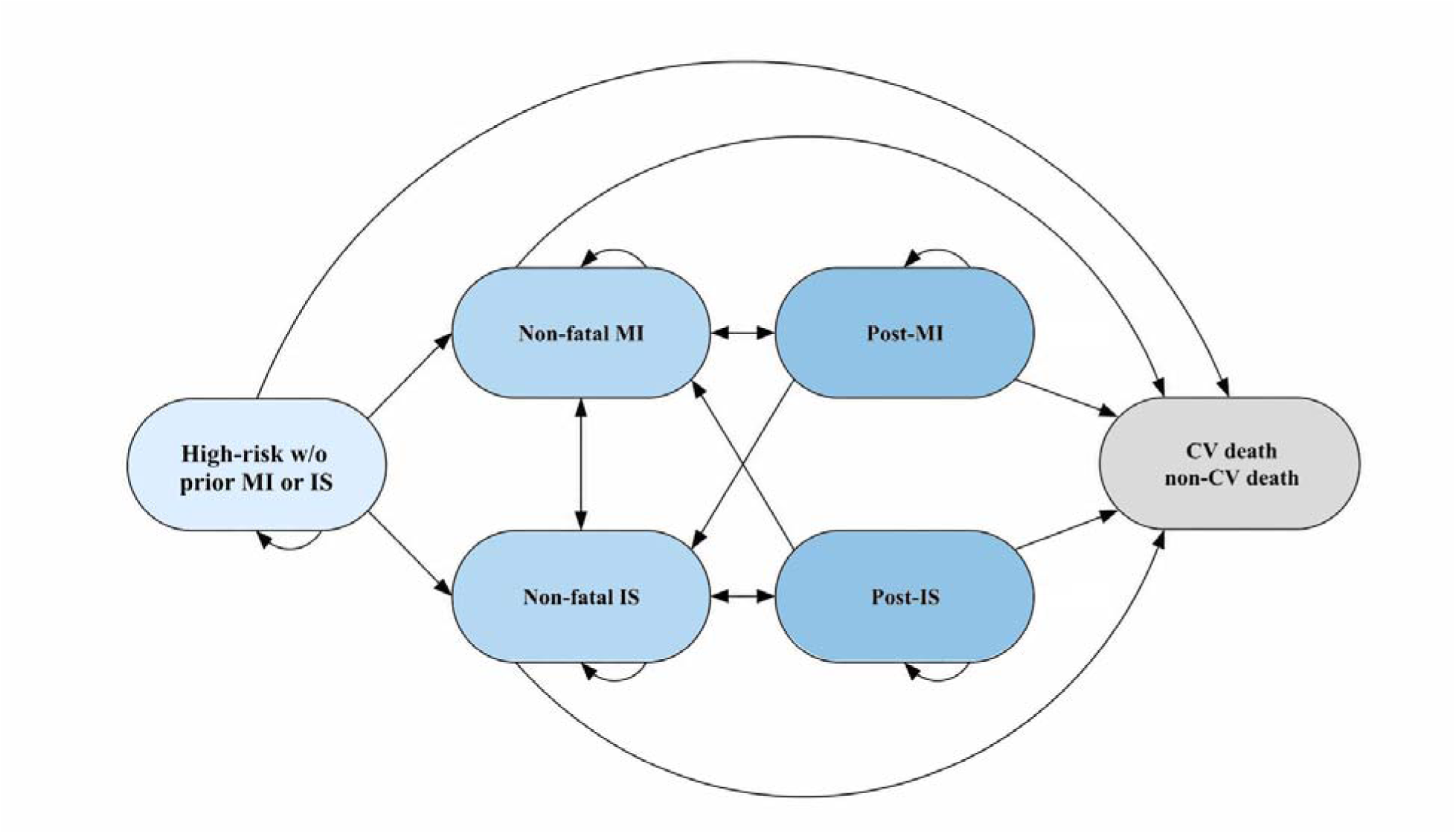
Markov Cohort State-Transition Model Diagram. CV, cardiovascular; IS, ischemic stroke; MI, myocardial infarction; w/o, without.

In the base case, a US payer perspective was considered. CV baseline risk and CV risk reduction before a first MI or IS were informed by VESALIUS-CV. After a non-fatal MI or IS, patients were assumed to transition to a recurrent ASCVD risk profile, and therefore, post-event CV baseline risk and CV risk reduction were informed by the FOURIER and FOURIER-OLE trials. CV risk reduction for evolocumab added to standard therapy compared with standard therapy alone was expressed as rate ratios per 38.67 mg/dL (1 mmol/L) reduction in LDL-C, derived from individual endpoint hazard ratios for MI, IS, CV death, and RV procedures from the evolocumab CV outcomes trials. Hazard ratios from VESALIUS-CV,^11^ FOURIER,^9^ and FOURIER-OLE^10^ trials were converted to rate ratios per 38.67 mg/dL using the following formula: Rate ratio per 38.67 mg/dL = hazard ratio^(1/[absolute LDL-C reduction/38.67]).

Model outcomes included major adverse cardiovascular events (MACE, defined as MI, IS, CV death), RV procedures, total costs, life-years (LYs), QALYs, and ICER. Cost-effectiveness was assessed using a willingness-to-pay (WTP) threshold of $120,000 per QALY gained, selected based on the 2025 AHA/ACC Statement on Cost/Value Methodology in Clinical Practice Guidelines.^16^ The current annual direct-to-patient price of evolocumab ($3,107 per year) was used as the drug acquisition cost input.^17,18^ Costs from the original Fonarow 2017 model were inflated to 2025 US dollars using the medical care component of the Consumer Price Index, where applicable.^19^ Key model inputs are summarized in Table 1.

**Table 1.**
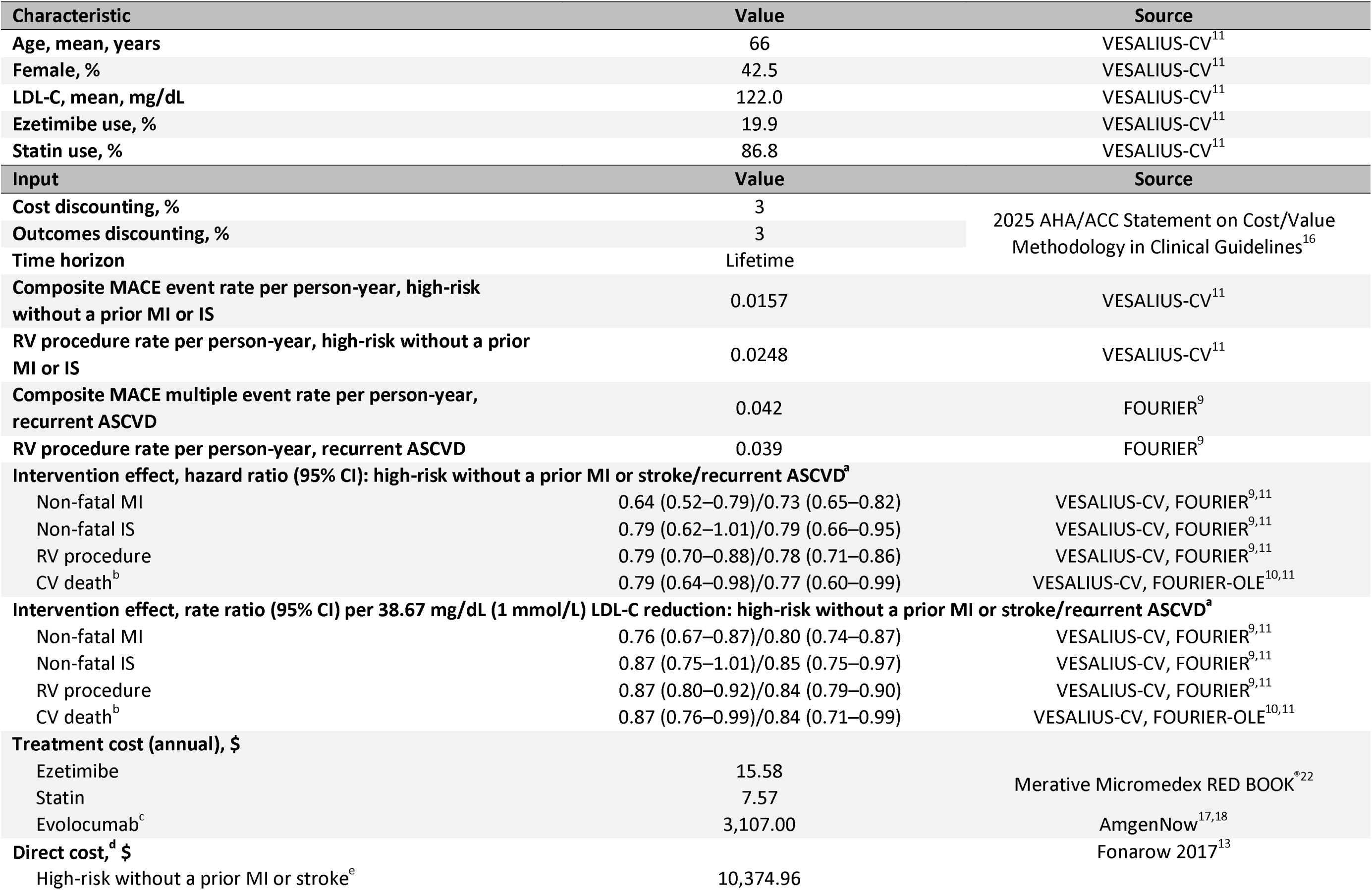
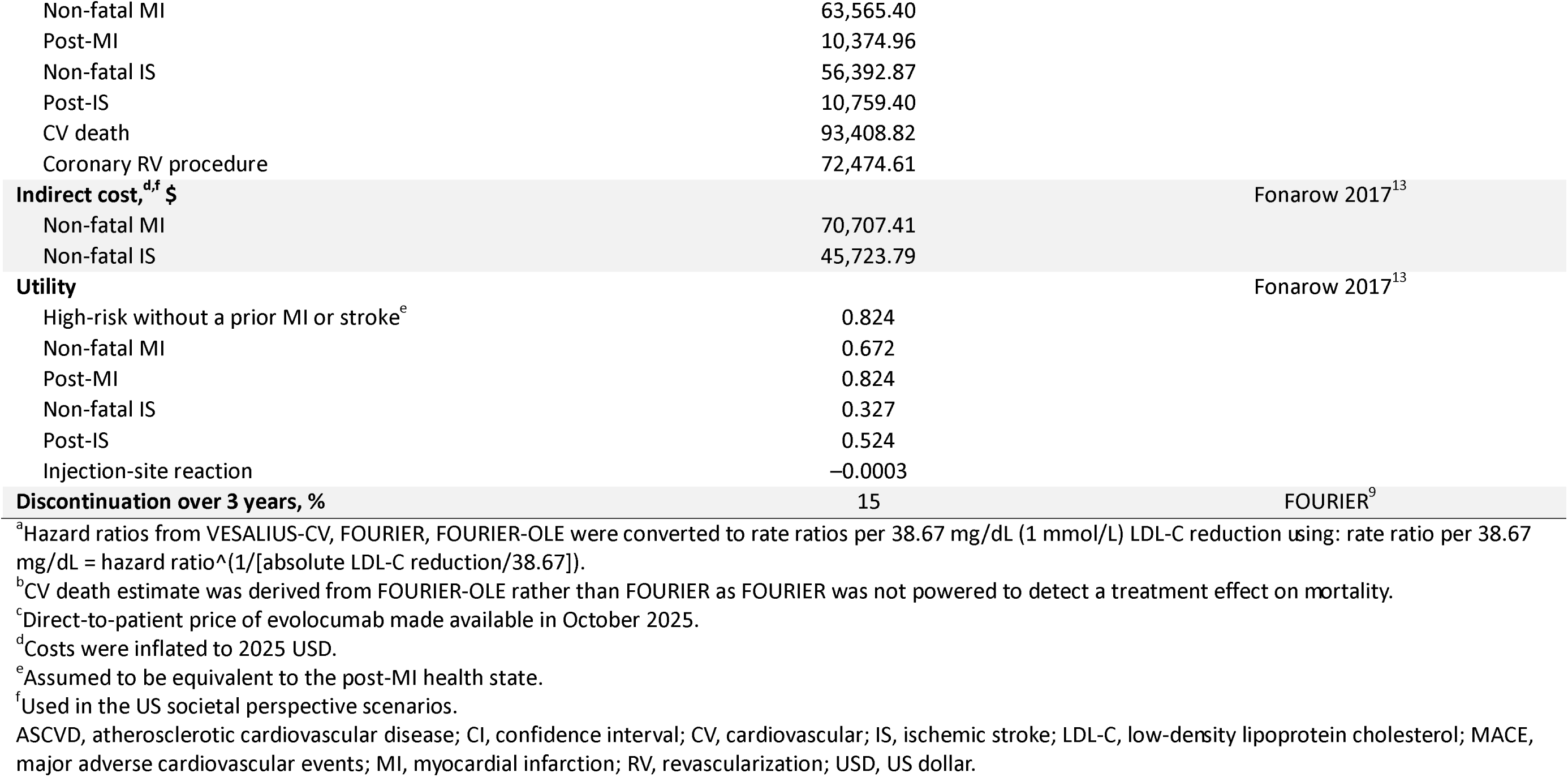
Patient Characteristics and Key Model Inputs.

A deterministic sensitivity analysis (DSA) was conducted by varying model parameters individually across plausible values. Additionally, three scenario analyses were conducted. Scenario 1 used the US payer perspective and applied CV risk reduction inputs from the Cholesterol Treatment Trialists’ (CTT) Collaboration 2010 meta-analysis rate ratios per 38.67 mg/dL (1 mmol/L) reduction in LDL-C.^20^ Scenarios 2 and 3 incorporated a US societal perspective using evolocumab CV outcomes trial-based and CTT Collaboration 2010 meta-analysis–based CV risk reduction estimates, respectively. The societal perspective differed from the payer perspective by including indirect costs associated with MI and IS events in addition to direct medical costs.

## Results

In the base-case analysis, evolocumab added to standard therapy was associated with both increased costs and improved outcomes compared with standard therapy alone in patients at high CV risk without a prior MI or stroke. At the current direct-to-patient cost of $3,107 per year and with CV risk reduction estimates from the evolocumab CV outcomes trials, evolocumab was associated with reductions of 0.24 MACE and 0.17 RV procedures per person over a lifetime horizon. Incremental costs were $24,430, with gains of 0.35 LYs and 0.34 QALYs. The corresponding ICER was $71,162 per QALY gained (Table 2).

**Table 2.** Cost-effectiveness Results for Base-Case and Scenario Analyses.

| Analysis | Event rates<sup>a</sup> | | Cost, \$ | | LY | | QALY | | ICER, \$/QALY | Estimated VBP,<sup>b</sup> \$ |
| --- | --- | --- | --- | --- | --- | --- | --- | --- | --- | --- |
|  | MACE | RV | Total | Δ | Total | Δ | Total | Δ |  |  |
| **<u>Base case:</u> US payer perspective, evolocumab CV outcomes efficacy data** |  |  |  |  |  |  |  |  |  |  |
| Standard therapy alone | 0.77 | 0.84 | 214,223 |  | 13.96 |  | 11.21 |  |  |  |
| Evolocumab plus standard therapy | 0.53 | 0.67 | 238,653 | 24,430 | 14.31 | 0.35 | 11.55 | 0.34 | 71,162 | 4,474 |
| **<u>Scenario 1:</u> US payer perspective, CTT Collaboration 2010 meta-analysis rate ratio per 38.67 mg/dL (1 mmol/L) LDL-C reduction** |  |  |  |  |  |  |  |  |  |  |
| Standard therapy alone | 0.77 | 0.84 | 214,223 |  | 13.96 |  | 11.21 |  |  |  |
| Evolocumab plus standard therapy | 0.48 | 0.54 | 230,730 | 16,506 | 14.33 | 0.37 | 11.60 | 0.39 | 42,094 | 5,594 |
| **<u>Scenario 2:</u> US societal perspective, evolocumab CV outcomes efficacy data** |  |  |  |  |  |  |  |  |  |  |
| Standard therapy alone | 0.77 | 0.84 | 234,837 |  | 13.96 |  | 11.21 |  |  |  |
| Evolocumab plus standard therapy | 0.53 | 0.67 | 252,400 | 17,563 | 14.31 | 0.35 | 11.55 | 0.34 | 51,160 | 5,033 |
| **<u>Scenario 3:</u> US societal perspective, CTT Collaboration 2010 meta-analysis rate ratio per 38.67 mg/dL (1 mmol/L) LDL-C reduction** |  |  |  |  |  |  |  |  |  |  |
| Standard therapy alone | 0.77 | 0.84 | 234,837 |  | 13.96 |  | 11.21 |  |  |  |
| Evolocumab plus standard therapy | 0.48 | 0.54 | 242,740 | 7,902 | 14.33 | 0.37 | 11.60 | 0.39 | 20,152 | 6,294 |

<sup>a</sup>Event rates are reported as events per person over the model-estimated survival time in each treatment arm: 20.3 years for evolocumab added to standard therapy and 19.7 years for standard therapy alone.
<sup>b</sup>Estimated at a willingness-to-pay threshold of \$120,000 per QALY gained, recommended by the 2025 AHA/ACC Statement on Cost/Value Methodology in Clinical Practice Guidelines. Values represent the model-estimated theoretical maximum net price for evolocumab at which the ICER would remain under this threshold. Estimates do not represent actual evolocumab prices in the marketplace.
ACC, American College of Cardiology; AHA, American Heart Association; CTT, Cholesterol Treatment Trialists'; CV, cardiovascular; ICER, incremental cost-effectiveness ratio; LDL-C, low-density lipoprotein cholesterol; LY, life-year; MACE, major adverse cardiovascular events; QALY, quality-adjusted life-year; RV, revascularization; VBP, value-based price.

DSA results showed that the ICERs remained below the WTP threshold across all parameters evaluated. Results for the 10 most influential parameters are presented in a tornado diagram (Figure 2). The model was most sensitive to individual event-specific risk reduction estimates from VESALIUS-CV.

**Figure 2.**
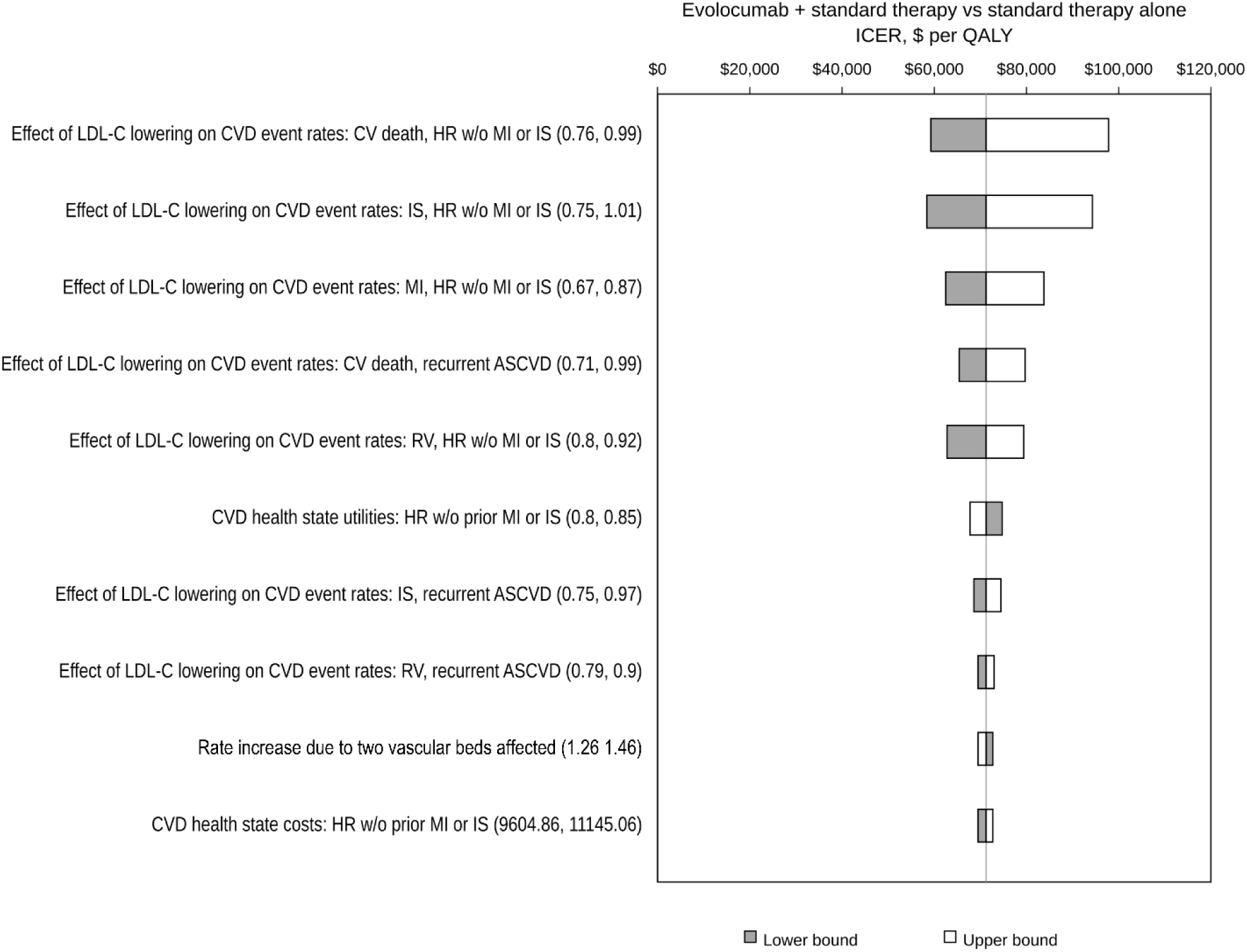
Tornado Diagram Showing Deterministic Sensitivity Analysis of Incremental Cost-Effectiveness Ratio. ASCVD, atherosclerotic cardiovascular disease; CV, cardiovascular; CVD, cardiovascular disease; HR, high-risk; ICER, incremental cost-effectiveness ratio; IS, ischemic stroke; LDL-C, low-density lipoprotein cholesterol; MI, myocardial infarction; QALY, quality-adjusted life-year; RV, revascularization; w/o, without.

In Scenario 1, using a US payer perspective and CTT Collaboration 2010 meta-analysis rate ratios,^20^ evolocumab was associated with greater lifetime event reductions, with 0.29 MACE and 0.30 RV procedures per person. Incremental costs were $16,506, with gains of 0.37 LYs and 0.39 QALYs, yielding an ICER of $42,094 per QALY gained.

Scenarios 2 and 3 adopted a US societal perspective while retaining the efficacy assumptions of the base case and Scenario 1, respectively. Accordingly, clinical outcomes, including MACE, RV procedures, LYs, and QALYs, were unchanged from their corresponding payer-perspective analyses.

Differences were noted in cost-based results, including incremental costs and ICERs, due to the inclusion of indirect costs. Incremental costs were $17,563 in Scenario 2 and $7,902 in Scenario 3, yielding ICERs of $51,160 and $20,152 per QALY, respectively.

## Discussion

This analysis evaluated the cost-effectiveness of adding evolocumab to standard therapy, compared with standard therapy alone, in patients with high CV risk but without a prior MI or stroke, modeled based on the recent VESALIUS-CV trial population. In the base-case and scenario analyses, the ICERs were substantially below the AHA/ACC-recommended WTP threshold of $120,000 per QALY gained, indicating that evolocumab is cost-effective at the current direct-to-patient price. This price was selected because it is publicly available and transparent,^17,18^ whereas prior analyses relied on list prices at the time or assumptions on discounts, as net prices were not publicly available.^13,14^ Since list prices generally exceed actual net prices after discounts and rebates, the direct-to-patient price may provide a pragmatic proxy for contemporary net prices.

Previous economic evaluations of evolocumab have largely focused on patients with clinically evident ASCVD.^13,14^ With the recent availability of the VESALIUS-CV results, this analysis extends prior economic evidence to a distinct care context focused on the prevention of a first major CV event in a high-risk population without prior MI or stroke. Although absolute event rates are generally lower in this population than in secondary prevention populations, VESALIUS-CV included patients with atherosclerosis or diabetes who remained at high risk for CV events. In this context, modeled reductions in MACE and RV procedures translated into ICERs substantially below the specified WTP threshold.

Taken together, the VESALIUS-CV trial findings and the results of the current analysis underscore the importance of earlier and more intensive LDL-C lowering in patients at high risk before the occurrence of a first major CV event. The risk reductions in 3-point MACE (hazard ratio: 0.75; 95% confidence interval [CI], 0.65–0.86) and 4-point MACE (hazard ratio: 0.81; 95% CI, 0.73–0.89) in VESALIUS-CV,^11^ combined with guideline support for PCSK9 monoclonal antibodies in select high-risk primary prevention populations, highlight an opportunity to improve implementation of evidence-based lipid-lowering therapy for patients who remain above LDL-C goals and at high CV risk despite standard care. Based on the current analysis, evolocumab may help address persistent treatment gaps by reducing lifetime CV events, preserving quality of life, and providing substantial economic value when used in appropriate high-risk patient populations.

A key strength of the current model is its ability to capture both the value of preventing first major CV events and longer-term consequences of disease progression by incorporating data from VESALIUS-CV, FOURIER, and FOURIER-OLE. Findings were consistent and yielded more favorable ICERs under alternative CV risk reduction inputs using the CTT Collaboration 2010 rate ratios per 38.67 mg/dL (1 mmol/L) LDL-C reduction. Additionally, the US societal perspective scenarios captured a more comprehensive estimate of the burden of MI and IS by accounting for event-related indirect costs, further improving cost-effectiveness estimates.

## Limitations

As with all model-based economic evaluations, findings should be interpreted within the context of the model structure, inputs, and underlying assumptions. In addition, clinical trial–derived inputs may not fully reflect patients treated in routine clinical practice due to differences in baseline risk, adherence, background therapy, and care patterns. Although real-world data specific to the VESALIUS-CV population are not yet available, evidence from clinically evident ASCVD suggests that routine-practice populations may have higher CV risk than trial populations.^21^ If a similar pattern applies here, use of trial-based baseline risk may have underestimated the absolute clinical benefit of evolocumab. However, the use of VESALIUS-CV, FOURIER, and FOURIER-OLE CV risk reduction estimates allowed the model to draw on relevant evolocumab outcomes evidence across high-risk and clinically evident ASCVD health states, and sensitivity and scenario analyses supported the robustness of the findings.

## Conclusion

These results suggest that evolocumab added to standard therapy was projected to improve health outcomes and to be cost-effective for patients at high CV risk without a prior MI or stroke. By extending cost-effectiveness evidence from a clinically evident ASCVD population to a high-risk population without prior MI or stroke, this analysis supports the potential value of earlier intensification of lipid-lowering therapy with evolocumab along the CV risk continuum to reduce future CV events and improve long-term health outcomes.

## Data Availability

All data produced in the present study are available upon reasonable request to the authors

## Acknowledgements

Medical writing and editorial support were provided by James Paterson, PhD, of Red Nucleus and funded by Amgen Inc.

